# Severity of lumbar spinal stenosis does not impact responsiveness to exercise-based rehabilitation

**DOI:** 10.1101/2024.09.20.24314088

**Authors:** Bahar Shahidi, Armin Zavareh, Connor Richards, Lissa Taitano, Kamshad Raiszadeh

## Abstract

Spine pain is a prevalent and costly condition affecting up to 85% of individuals throughout their lifetime, and spinal stenosis is one of the most debilitating sources of spine pain. Although conservative management is the first line of treatment for spinal stenosis, severe cases often are directly referred to surgical intervention due to the belief that conservative strategies delay necessary treatment. However, there are no studies supporting the premise that individuals with more severe stenosis respond poorly to conservative management. The purpose of this study was to compare improvements in pain, disability, strength, medication usage, and patient goals in response to an exercise-based physical therapy program across 1,806 individuals with mild, moderate, or severe lumbar spine stenosis. Participants demonstrated significant improvements in all variables of interest (p<0.001), and 11.5% of participants reported cessation of narcotic use with treatment. There were no significant differences in treatment response across mild, moderate, or severe stenosis groups for any outcome (p>0.546). Exercise-based rehabilitation is as beneficial in the short term for individuals presenting for nonoperative care with severe stenosis compared to their milder counterparts. Future research is needed to evaluate long term durability and cost effectiveness of rehabilitation in this patient population.

## INTRODUCTION

Low back pain (LBP) is a highly prevalent musculoskeletal condition that is responsible for the greatest number of years lived with disability in the general population[1-6]. Between 65-85% of individuals experience LBP at some point in their life [7]. The presence of symptomatic lumbar spinal stenosis has a strong negative influence on health related quality of life[8]. Of symptomatic spinal pathologies, spinal stenosis is one of the most common, costly, and difficult to remediate. Data on the natural history of stenosis is mixed, with some studies reporting favorable short-term (<1 year) outcomes in 50% of patients[9], and others report that up to 60% experience symptom deterioration over time[10]. Further, longer term outcomes indicate that a significant proportion (between 15-30%) of individuals with stenosis experience worsening of stenosis[11, 12].

Initial conservative management of spinal stenosis is typically non-operative. However, conservative management for stenosis is not standard worldwide, and often moderate and severe cases are streamlined to more immediate surgical intervention[13, 14]. The literature on efficacy of surgical management as compared to conservative management is conflicting; some studies report that surgical management is superior to conservative management[15-17], however these studies also report high incidence of side effects (10-30%) and most have inclusion criteria requiring eligible participants to have already failed an initial trial of 3-6 months of conservative management[18], potentially biasing results in favor of surgical management. On the other hand, some studies demonstrate that conservative management is equivalent to surgical treatment[18, 19], or is even superior in mild cases[20]. Given these conflicting reports, reviews of the literature evaluating the comparative efficacy of conservative versus surgical management of spinal stenosis state that the quality of the literature is too poor to allow for clear clinical recommendations to be made[18].

One factor confounding these investigations is that conservative management is broadly defined. Individual or combined applications of pharmacological management, physical therapy, injections, and education are often discussed interchangeably under the umbrella of “conservative treatment” when comparing to surgical management. However, the individual components of these treatments have different physiological targets, dosing regimens, and different treatment effects. Of the common conservative treatments, exercise-based rehabilitation has shown to demonstrate consistent efficacy; with studies reporting that a large proportion of patients with stenosis participating in a physical therapy program with an exercise component experience short and long term symptom improvement (up to 3 years)[21-23], and have reduced incidence of surgery^9^. However, some evidence suggests that these improvements are not observed when the level of radiographic stenosis is considered severe[22, 24, 25]. Indeed, some proposed clinical decision-making algorithms recommend that individuals with severe stenosis should not be considered for physical therapy-based treatment at all and should be prioritized for surgical intervention in order to reduce further disease progression[13, 14]. To date, very little evidence exists that supports or refutes the differential efficacy of exercise-based rehabilitation programs across individuals with varying levels of spinal stenosis. Therefore, the purpose of this investigation was to evaluate whether improvements in pain, disability, strength, medication usage, and patient goals in response to an exercise-based physical therapy program differed across individuals with mild, moderate, or severe lumbar spine stenosis. Our hypothesis was that individuals with severe stenosis would demonstrate significantly smaller improvements in outcomes as compared to those with mild or moderate stenosis.

## METHODS

### Participants

This was a secondary analysis of data from a clinical trial registry (NCT #04081896) approved by the local ethical review board (WIRB #1252493). All participants provided informed consent to participate in this study. Participants were included if they were 1) prescribed exercise-based rehabilitation to address their spinal condition 2) had radiographic imaging within 6 months of initiation of their physical therapy program, 3) that imaging indicated stenosis at one or multiple levels in the lumbar spine, and 4) they had complete baseline pain and disability data. Participants were required to have undergone at least 3 treatment sessions to evaluate the influence of the program on patient outcomes, and they must have completed their prescribed program within 6 months of an initial evaluation.

### Exercise-based rehabilitation program

The exercise-based physical therapy program was administered in an integrated practice unit (IPU) consisting of a multidisciplinary treatment team of physical therapists, orthopedic spine surgeons, spine-trained physician assistants, and pain specialist consultants. The treatments were implemented under direct supervision of a licensed physical therapist or physical therapist assistant, and included machine-based resistance exercises prescribed and progressed as previously described in detail^2^ along with directional preference exercises, and patient education on sleep, nutrition, posture, and ergonomics as needed based on impairments identified upon initial physical therapy evaluation[26].

### Outcomes measured

The primary predictor variable of interest was stenosis severity, categorized as mild, moderate, or severe based on radiology reports from imaging obtained prior to participation in the physical therapy program. For individuals with multiple levels of stenosis, they were categorized according to the spinal level with the most severe designation. Primary outcomes included changes in pain, disability, medication use, patient goal achievement, and paraspinal strength. Pain was measured using a visual analogue scale from 0-100 points and was collected for the back and legs as appropriate[27]. Because leg pain was collected bilaterally, the maximum value was used across the left and right leg as the representative leg pain value. Functional status was measured using the Oswestry Disability Index (ODI) [28]. The use of narcotic medications for pain management was categorized according to frequency of use (none, <1/day, 1-2/day, 3-5/day, and >6/day). Patient goals were measured using the Patient Specific Functional Scale (PSFS), which is a 0-10 scale indicating the patient’s ability to achieve self-entered goals, with higher scores indicating greater goal achievement[29]. Strength of the lumbar paraspinal muscles was measured for resisted extension using a MedX isokinetic dynamometer (MedX Holdings Inc., Cheyenne WI). Response to the program was measured as the change in outcomes from baseline to the time of discharge. Covariates included demographic, stenosis-specific, and program-specific variables. Demographic variables included age, gender, and body mass index (BMI). Stenosis-specific variables included stenosis type (central or foraminal). Program-specific variables included the number of days in the program and number of visits attended.

### Statistical Analysis

Each measurement was compared across stenosis severity groups using a one-way ANOVA with Sidak post-hoc corrections for multiple comparisons for continuous variables, and chi-square analysis for categorical or binary variables. Data was confirmed for normality of distribution. Both intention-to-treat based analysis and as-treated analysis approaches were used to evaluate outcomes. For intention to treat analyses, any missing outcomes were assumed to have not changed from the baseline. In the as-treated analysis, only complete data was analyzed for a given outcome. Secondary covariates that were significantly different across stenosis severity groups were included in a multivariate linear regression model (for continuous outcomes) or logistic regression model (for categorical outcomes) along with the primary predictor of interest to adjust for confounders. Adjusted p-values of <0.05 were considered statistically significant.

## RESULTS

### Participants

An initial sample of 1,806 individuals initiated an exercise-based rehabilitation program for their spine condition. Of those, 300 participants were excluded due to completing less than 3 visits, and 152 participants were excluded due to breaks in care resulting in more than 6 months treatment duration. Another 18 did not have baseline data for the outcomes of interest. There were 1,336 (74%) participants resulting for analysis (Figure 1). Individuals with mild stenosis represented 19.3% of the cohort, those with moderate stenosis represented 36.6% of the cohort, and those with severe stenosis represented 44.1% of the cohort. The majority of participants were in their 6^th^ decade of life, were overweight, and were female. Individuals with severe stenosis were significantly older (p<0.001), heavier (p=0.040), and more male (p=0.024) than those with mild stenosis. Of the 1,336 participants who initiated the program, all participants (100%) had complete pain and medication use outcomes data at the time of discharge, and 64.0% had both pain and disability data upon discharge. Individuals who did not have disability data upon discharge still participated fully in the program but did not complete the disability questionnaire. At baseline, participants reported moderate levels of pain and disability, low-moderate ability to achieve goals, and had chronic (>3 months duration) symptoms. There was a trend for individuals with severe stenosis to have longer symptom durations and greater disability levels than those with mild or moderate stenosis for the back (p=0.051 and 0.072 respectively). Most of the stenosis was classified as central, with central stenosis being more prevalent in those with severe stenosis as compared to mild or moderate (p<0.001). Duration in the program averaged approximately 13 visits over the course of 92 days, with no differences in attendance across stenosis severity types (p>0.092). Baseline characteristics across stenosis severity groups can be found in Table 1.

**Table 1.** Baseline characteristics of participants. Values are represented as mean (standard deviation) or percent. Significant p-values are indicated in bold. BMI=Body Mass Index; ODI=Oswestry Disability Index; PSFS=Patient Specific Functional Scale

| Variable | N | Mild | Moderate | Severe | p-value |
| --- | --- | --- | --- | --- | --- |
| Age | 1336 | 57.12 (15.07) | 64.76 (14.70) | 71.19 (11.48) | **<0.001** |
| BMI | 1333 | 27.05 (5.48) | 28.01 (5.79) | 28.05 (5.50) | **0.040** |
| Gender (% female) | 1336 | 53.5% | 55.0% | 47.0% | **0.024** |
| Days in program | 1336 | 90.80(35.44) | 91.98 (38.23) | 92.39 (36.84) | 0.848 |
| Number of Visits | 1336 | 12.47 (6.62) | 13.40 (7.00) | 13.58(6.93) | 0.092 |
| Symptom duration<br>(days) | 1282 | 73.38(112.12) | 74.68 (105.81) | 90.23 (127.26) | 0.051 |
| Stenosis Type (%<br>central) | 1336 | 41.1% | 53.4% | 65.4% | **<0.001** |
| ODI initial | 1787 | 30.62(14.59) | 32.62 (15.07) | 33.16 (14.97) | 0.072 |
| PSFS initial | 1322 | 3.13(2.08) | 3.17 (2.02) | 3.14(2.18) | 0.947 |
| Pain | 1336 | 57.29 (18.65) | 58.58 (20.84) | 60.31 (21.23) | 0.116 |
| Lumbar Extensor<br>Strength | 1069 | 142.53 (80.76) | 132.67 (76.29) | 137.64 (80.13) | 0.321 |
| Narcotic Medication<br>Use (%) | 1336 |  |  |  | 0.734 |
| None | 1029 | 20.0% | 36.5% | 43.4% |  |
| <1/day | 111 | 18.0% | 37.8% | 44.1% |  |
| 1-2/day | 110 | 19.1% | 31.8% | 49.1% |  |
| 3-5/day | 74 | 12.2% | 40.5% | 47.3% |  |
| >6/day | 12 | 16.7% | 50.0% | 33.3% |  |

**Figure 1.**
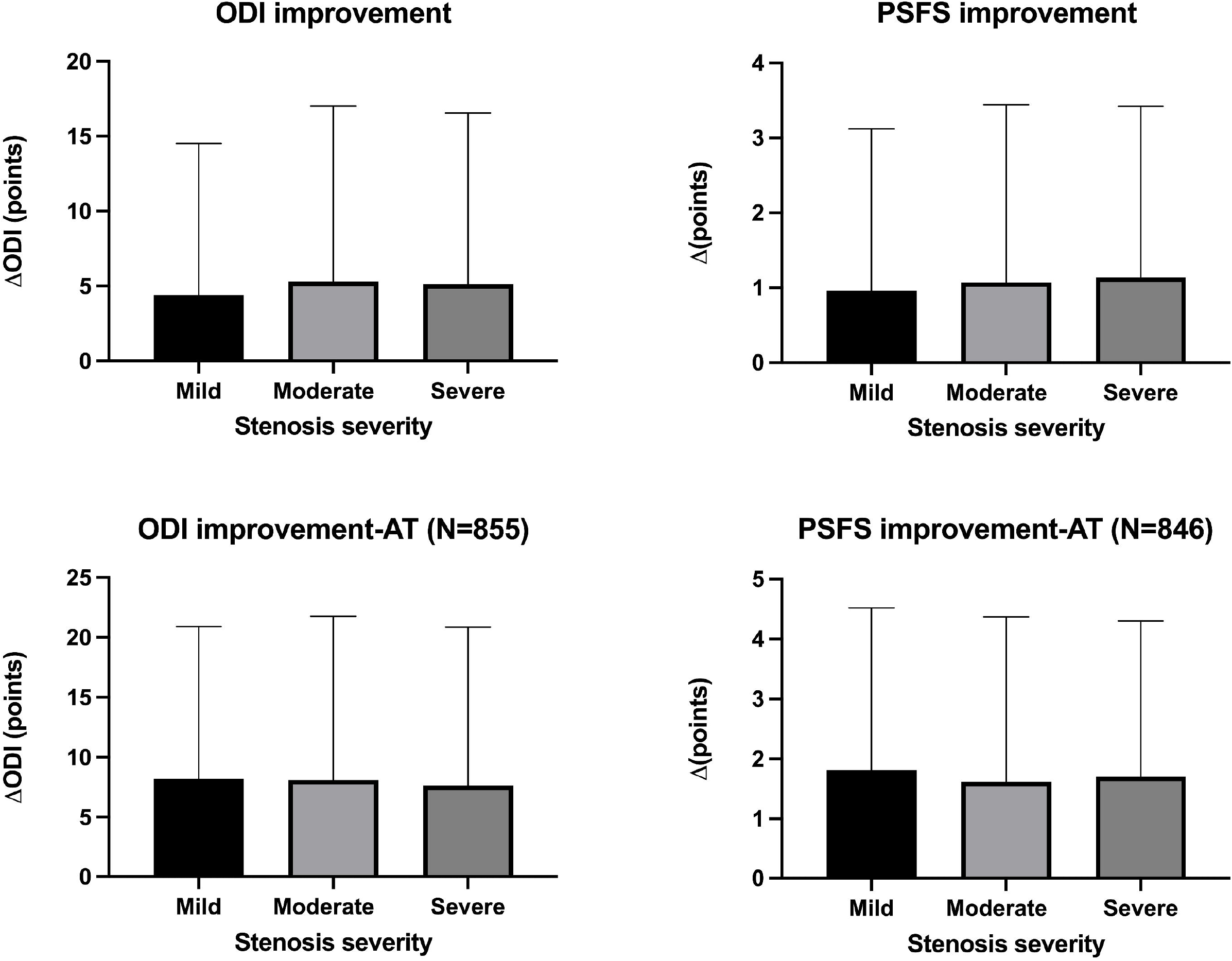
Improvements in disability and goal achievement scores across groups for the intention to treat analysis (top row) and the as-treated(AT) analysis (bottom row). Data are represented as mean and standard deviation.

### Intention to treat analysis

For the intention-to-treat outcomes, the average improvement in ODI was 5.1(11.3) points (p<0.001). Average improvement in goal achievement (PSFS score) was 1.7 (2.7) points (p<0.001; **Figure 1**). Average improvement in pain was 26.6(24.3) points (p<0.001; **Figure 2**)). There were no significant differences in the magnitude of improvement reported for disability, goal achievement, or pain across stenosis severity groups (p> 0.49). The frequency of narcotics use was reduced in 15.6% of patients, with 11.5% ceasing use of narcotics altogether (**Figure 3**). There was no significant difference in narcotic use reduction across groups (p=0.72). Participants improved their lumbar extension strength an average of 35.1(66.1)% (p<0.001). Individuals with moderate stenosis demonstrated larger improvements in back extension strength in response to treatment as compared to those with mild stenosis (p=0.066; **Figure 4)**. Adjusting for significant covariates of age, BMI, gender, and stenosis type did not change the results.

**Figure 2.**
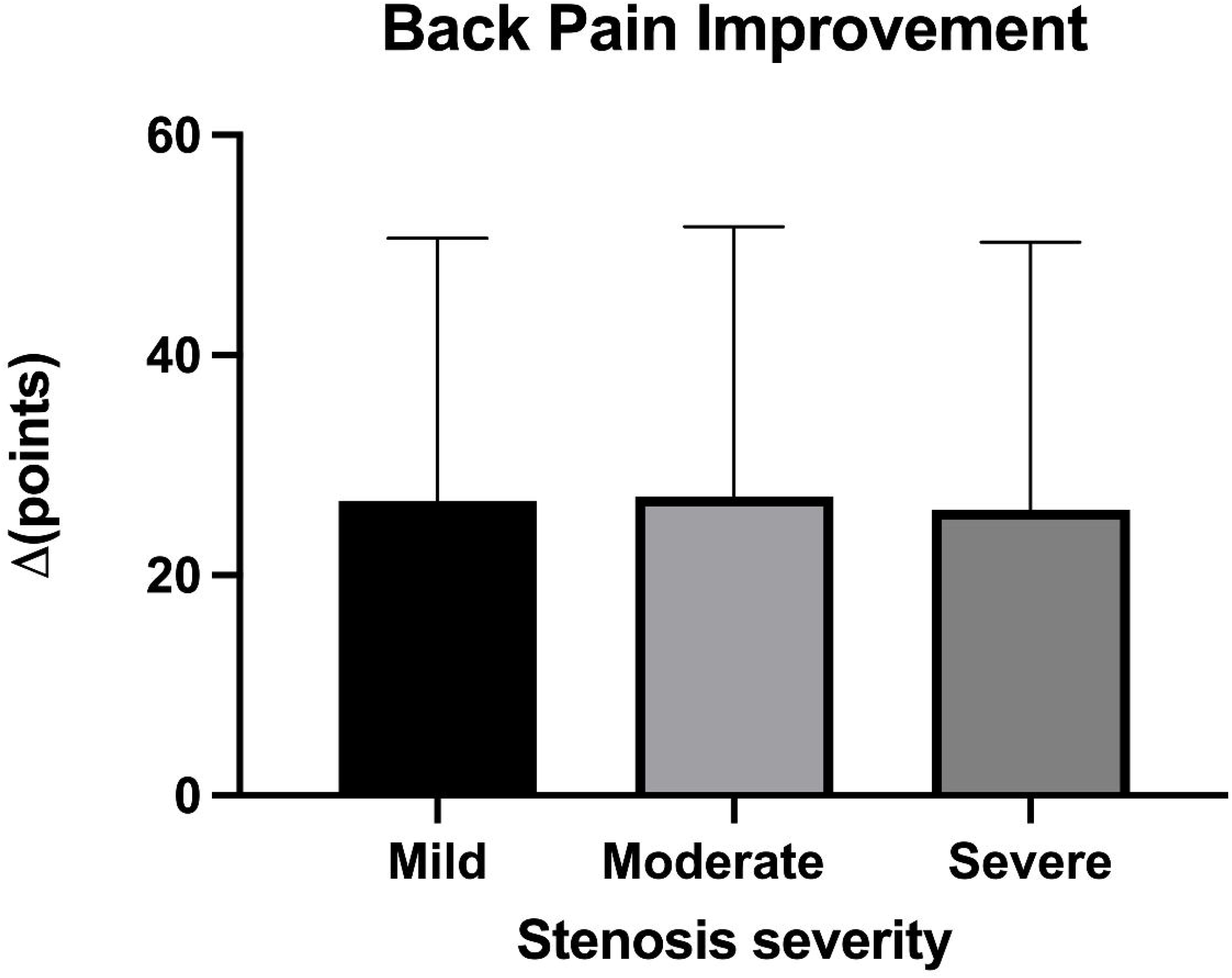
Improvements in back pain for all participants across stenosis severity groups. Data are represented as mean and standard deviation.

**Figure 3.**
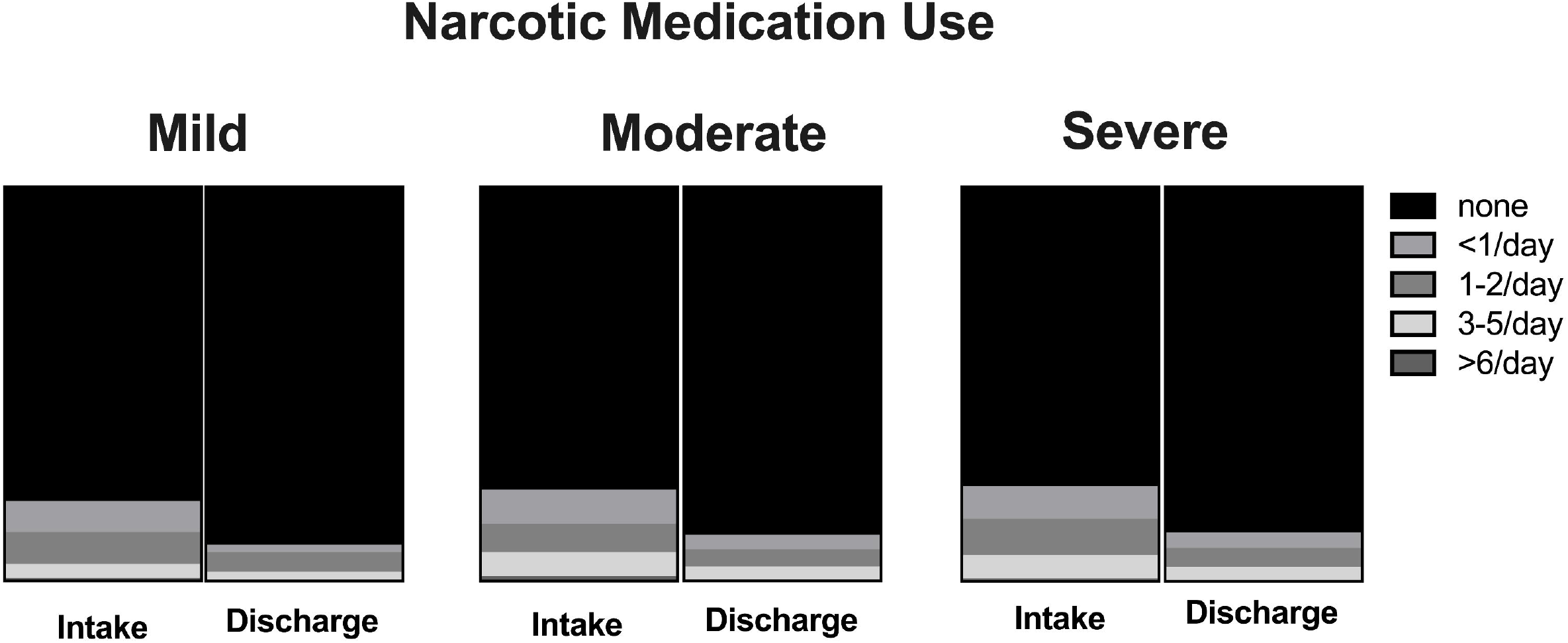
Changes in frequency of narcotic medication use across mild, moderate, and severe stenosis categories.

**Figure 4.**
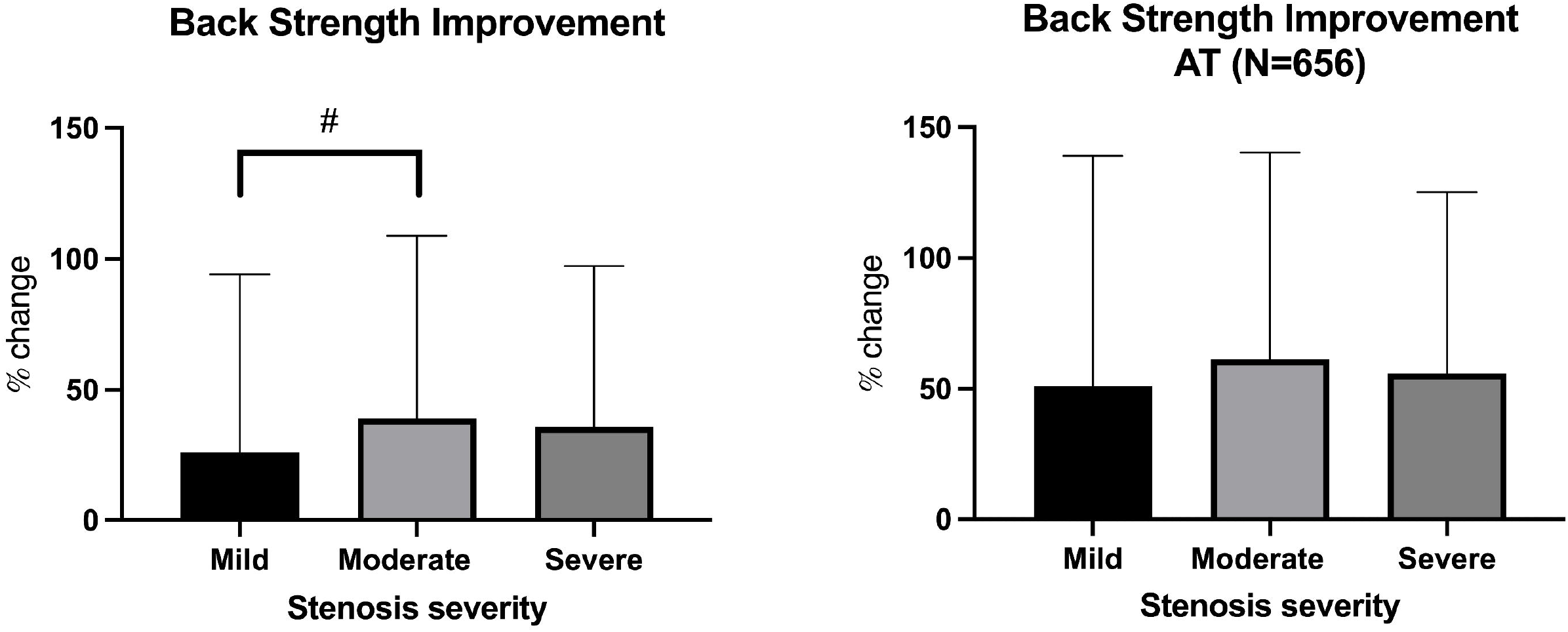
Improvements in back extensor strength as measured by an isokinetic dynamometer for the intention to treat(left) analysis and as-treated(AT;right) analysis. Data are represented as mean and standard deviation. # indicates a trend toward a difference at p<0.1).

### As-treated analysis

Due to lack of complete data for some outcomes, disability, goal achievement, and strength outcomes were also included in an as-treated analysis. Complete data was available for 64.0% of patients for disability, 63.3% of patients for PSFS scores, and 49.1% of patients for strength scores. The average improvement in ODI was 7.9(13.3) points (p<0.001), and improvement in goal achievement (PSFS score) was 1.7(2.7) points (p<0.001). There were no significant differences in the magnitude of improvement reported for disability or goal achievement across stenosis severity groups (p> 0.769; **Figure 1**). Lumbar paraspinal strength improved by an average of 57.1(76.5)%, and there were no differences in magnitude of strength improvement across stenosis severity groups in the as-treated analysis (p=0.471; **Figure 4**).

## DISCUSSION

This study demonstrated that individuals with varying levels of stenosis severity demonstrated similar, and statistically significant improvements in pain, disability, goal achievement, and spinal extensor strength, along with reductions in narcotic medication use in response to an exercise-based interdisciplinary rehabilitation program. These improvements were clinically significant (reached minimal clinically important difference thresholds) for pain, but not disability or goal achievement regardless of the analysis approach (intention to treat vs. as-treated). We found a trend for larger improvements in lumbar extensor strength in individuals with moderate lumbar spine stenosis as compared to those with mild stenosis in the ITT analysis, but not the ATT analysis. These findings are contrary to our hypothesis that individuals with more severe stenosis would not be as responsive to an exercise-based physical therapy program as compared to those with mild or moderate disease.

Our results are consistent with previous literature demonstrating that interventions including exercise can be beneficial for improving pain and function in individuals with lumbar spine stenosis when compared to no treatment[21-23, 30], particularly in the short term. However, the radiographic severity of stenosis and its independent influence on potential for treatment response has not been explicitly investigated. More common investigations are those that compare responsiveness across varying symptom severities, and frequently report that individuals with more severe symptoms are more likely to require healthcare management beyond what is available with conservative management strategies[31]. Interestingly, our data did not suggest that more severe radiographic stenosis was associated with greater symptom severity or functional deficit; instead, demographic and structural characteristics such as age, gender, and stenosis type were related. Overall, the lack of relationship between structural features and symptom severity is in line with several previous reports acknowledging that changes in spinal anatomy as a function of age and gender are common observations, regardless of the presence of symptoms[32-34]. Indeed, the prevalence of spinal stenosis in imaging of asymptomatic older individuals, the frequently ambiguous and inconsistent symptoms, and the diverse nature of spinal stenosis all contribute to the complexity of its management.

Despite the improvements observed across all groups in the measured outcomes, many of these improvements did not reach clinical significance according to defined thresholds[35]. Their magnitudes are similar to previous studies using similar outcomes for pain and disability [18, 30] but demonstrated greater reductions in medication use[21]. Given the observation that a significant proportion of individuals reduced their analgesic medication intake in response to treatment, it is possible that the treatment effect for symptom reduction is confounded by this concurrent change. The optimal balance between pharmacological pain management and symptom resolution requires further elucidation, particularly considering the high prevalence of narcotic use and misuse in populations with chronic spine pain.

This study is not without limitations. First, our follow-up duration only spanned the duration of the treatment program (approximately 3 months) and as such is likely insufficient for assessing the durability of the observed effects over the long term. Second, our study design lacked a control group, which would have allowed for a comparative assessment of the natural history of spinal stenosis across different severities. However, given that the duration of symptoms was on average 6 months or more in our cohort, the likelihood that large variations in symptoms within the study period are unrelated to treatment are less likely. Our cohort was not balanced across groups according to some baseline characteristics, which may have led to confounding influences of age, gender, and stenosis type (central or foraminal) on interpretation of treatment outcomes. Although we attempted to mitigate this issue by using a multivariate analysis to adjust for these confounders, a more rigorous study design is necessary to address this limitation. Finally, this study may suffer from selection bias in that it is possible that individuals that had more severe symptoms either elected not to initiate this modality of conservative management at all, or were deemed by their referring provider to not be fit for conservative management. However, since many insurance providers will not approve surgical interventions until individuals have demonstrated failure in some form of conservative management, this may be a smaller concern. Future research endeavors should aim to investigate whether short-term enhancements in pain and functional capacity in individuals with severe stenosis are sustained over an extended duration, and whether exercise-based physical therapy can effectively forestall the necessity of long-term transition to more costly and invasive interventions such as surgery.

## CONCLUSIONS

Individuals with severe stenosis undergoing an exercise-based rehabilitation program demonstrate similar short-term improvements in spine pain, disability, goal achievement, strength, and narcotic use cessation when compared to those with mild or moderate stenosis who presented for nonoperative treatment to an integrated practice unit. Further research is needed to evaluate the durability of these improvements and if they influence the likelihood of progression to more invasive and costly interventions such as surgery in the long term.

## DATA AVAILABILITY

Data will be made available upon reasonable request to the corresponding author.

## AUTHOR CONTRIBUTIONS

BS and SZ wrote the main manuscript text, BS and CR performed data analysis, LT and KR generated patient data from the clinical trial and reviewed the manuscript. All authors approved the final manuscript.

